# Genomic sequencing surveillance of patients colonized with vancomycin-resistant *Enterococcus* (VRE) improves detection of hospital-associated transmission

**DOI:** 10.1101/2024.05.01.24306710

**Authors:** Alexander J. Sundermann, Vatsala Rangachar Srinivasa, Emma G. Mills, Marissa P. Griffith, Eric Evans, Jieshi Chen, Kady D. Waggle, Graham M. Snyder, Lora Lee Pless, Lee H. Harrison, Daria Van Tyne

**Author notes:** **Corresponding Author:** Daria Van Tyne; University of Pittsburgh BST E1059 200 Lothrop Street Pittsburgh, PA 15213. **Summary:** This study demonstrates that inclusion of VRE rectal swabs in hospital surveillance enhances detection of geotemporal pathogen transmission. Rectal swab sampling and forward VRE transmission was associated with readmission and death in 10-20% of VRE patients.

## Abstract

**Background:** Vancomycin-resistant enterococcal (VRE) infections pose significant challenges in healthcare. Transmission dynamics of VRE are complex, often involving patient colonization and subsequent transmission through various healthcare-associated vectors. We utilized a whole genome sequencing (WGS) surveillance program at our institution to better understand the contribution of clinical and colonizing isolates to VRE transmission.

**Methods:** We performed whole genome sequencing on 352 VRE clinical isolates collected over 34 months and 891 rectal screening isolates collected over a 9-month nested period, and used single nucleotide polymorphisms to assess relatedness. We then performed a geo-temporal transmission analysis considering both clinical and rectal screening isolates compared with clinical isolates alone, and calculated 30-day outcomes of patients.

**Results:** VRE rectal carriage constituted 87.3% of VRE acquisition, with an average monthly acquisition rate of 7.6 per 1000 patient days. We identified 185 genetically related clusters containing 2-42 isolates and encompassing 69.6% of all isolates in the dataset. The inclusion of rectal swab isolates increased the detection of clinical isolate clusters (from 53% to 67%, P<0.01). Geo-temporal analysis identified hotspot locations of VRE transmission. Patients with clinical VRE isolates that were closely related to previously sampled rectal swab isolates experienced 30-day ICU admission (17.5%), hospital readmission (9.2%), and death (13.3%).

**Conclusions:** Our findings describe the high burden of VRE transmission at our hospital and shed light on the importance of using WGS surveillance of both clinical and rectal screening isolates to better understand the transmission of this pathogen. This study highlights the potential utility of incorporating WGS surveillance of VRE into routine hospital practice for improving infection prevention and patient safety.

## INTRODUCTION

Vancomycin-resistant enterococci (VRE) are a major cause of healthcare-associated infections and are associated with high morbidity and mortality. Within the United States, VRE have been estimated to cause over 54,000 infections in hospitalized patients and over 5,400 deaths annually.^1^ VRE have the ability to persist in the hospital environment and colonize the gastrointestinal tracts of patients.^2–4^ Immunocompromised individuals and other high-risk patients with invasive procedures or indwelling devices are at higher risk for VRE infections.^5,6^ The limited treatment options that are active against VRE further complicates the management of these infections.^4^

The transmission dynamics of VRE within healthcare settings are complex and involve multiple factors. Patients may become colonized and harbor VRE in their gastrointestinal tract, thereby acting as a moving reservoir for transmission to other patients.^7^ Direct contact with contaminated surfaces, healthcare workers, or other colonized patients plays a significant role in the spread of VRE.^8^ Outbreaks of VRE in healthcare settings are well described in the literature including from medical procedures, environmental contamination on specialized units, or emergence and transmission within individual hospitals.^9–13^ Many of these outbreaks are characterized by prolonged spread over the course of several months or years.^14,15^

Traditional epidemiological methods are likely to be insufficient in elucidating the complex spread of VRE in healthcare settings. The limitations of these methods stem from their inability to differentiate between closely related strains, identify long-range transmission events, and accurately discern geo-temporal patterns of transmission.^16^ Consequently, a deeper understanding of the routes and mechanisms of VRE transmission in modern hospitals is crucial for effective infection control and the development of targeted interventions. The use of whole genome sequencing (WGS) has emerged as a useful tool to accurately differentiate the relatedness of pathogens, enabling investigators to confirm or refute the presence of transmission events or outbreaks.^16–18^ In the context of VRE, WGS has been invaluable in directing interventions and understanding transmission patterns.^19–21^ However, the majority of these studies use WGS in reaction to suspected outbreaks rather than as pro-active surveillance to identify and characterize undetected transmission patterns of endemic pathogens (like VRE) in healthcare settings.

In November 2016 we began the Enhanced Detection System for Healthcare-Associated Transmission (EDS-HAT) program at our institution, which combines WGS surveillance of major bacterial pathogens, including VRE, to better detect and intervene on healthcare outbreaks.^22^ The purpose of this study was to summarize the genomic epidemiology and transmission dynamics of VRE at our institution using WGS surveillance of clinical and rectal screening VRE isolates during a 34-month period.

## METHODS

### Study setting

The UPMC Presbyterian Hospital is an adult tertiary care hospital with over 750 beds (including 134 critical care beds) and performs over 400 solid organ transplants annually. During the study period contact precautions were used for the care of any patient with a history of VRE carriage. Patients with VRE carriage were cared for in single-bedded rooms or were cohorted in double-bedded rooms only if the roommate was known to have VRE carriage. No method for performing or evaluating for decolonization was employed. Ethics approval was obtained from the University of Pittsburgh Institutional Review Board (Protocol STUDY21040126).

### Isolate collection

We collected potentially healthcare-associated VRE clinical cultures, as defined by patients with a hospital stay ≥3 days or a recent healthcare exposure in the prior 30-days, from November 2016 through August 2019. From September 2017 through May 2018, we also collected and sequenced VRE isolates obtained through rectal swab screening of admitted patients. As part of standard infection prevention practice during the study period, patients admitted to our hospital without prior VRE history were screened via rectal swab on admission, weekly, and at discharge until positive. Monthly VRE incidence was calculated using patient days at our hospital with newly detected VRE from clinical or rectal swab cultures.

### Retrospective whole genome sequencing and analysis

Genomic DNA was extracted from VRE isolates that were grown overnight at 37 ^O^C on a blood agar plate. WGS was performed on the Illumina platform using 2 x 150bp paired end reads on a NextSeq2000. The resulting reads were assembled using SPAdes,^23^ and multilocus sequence types (STs) were identified using PubMLST.^24^ Isolate genomes were compared to one another using snippy (https://github.com/tseemann/snippy) and split kmer analysis (SKA).^25^ Clusters of genetically related isolates were identified using hierarchical clustering with average linkage and a cut-off of 20 single nucleotide polymorphisms (SNPs) as determined by SKA. Evolutionary rates were estimated by linear regression of pairwise SNPs calculated using snippy or SKA versus days between collection dates for all isolates sampled from the same patient.

### Transmission analysis

Transmission analysis was performed using a geo-temporal approach considering clustered patients who shared a hospital unit commonality using two separate approaches (**Figure S1**). One scenario considered the prior 100 days from the exposed patient’s positive culture date to have a unit commonality concurrently or after the clustered index patient, who was already culture-positive.^26^ The second scenario was similar but considered the 50 days after the exposed patient’s positive culture date for an index patient to have a positive culture date. This analysis was performed for both clinical and rectal isolates and for clinical isolates only to determine the contribution of rectal isolates in elucidating possible transmission routes for clinical isolates. We additionally calculated a transmission rate by summing the number of transmissions per day during the clinical-only time period versus the combined clinical and rectal time period.

### Outcome analysis

In order to estimate the impact of transmission from patients with VRE colonization, we recorded 30-day all-cause clinical outcomes including intensive care unit (ICU) admission, hospital readmission, and death among patients with a clinical isolate genetically related to an isolate from a patient with colonization. The subset of outcomes was defined as those that were directly associated with VRE infection. We additionally characterized whether each clinical isolate was classified as an HAI using National Healthcare Safety Network definitions, as adjudicated by infection preventionists as part of normal operations.^27,28^

### Statistical analysis

Mann Whitney U-test was used to assess differences in days between same-ST isolates versus different-ST isolates among patients that were repeatedly sampled. Liner regression was performed to calculate rates of SNP accumulation over time and how they differed between SKA versus snippy. Differences in average cluster size between clinical isolates only versus clinical and rectal swab isolates were assessed with a one-sided difference in means t-test. To calculate the difference in proportion of clustered isolates between clinical isolates only versus clinical and rectal swab isolates, a one-sided difference in proportion hypothesis test was performed. A Wilcoxon Signed Ranked test was used to assess differences in transmission rates between collection periods with clinical isolates only versus clinical and rectal swab isolates.

## RESULTS

### VRE incidence and carriage

From November 2016 to August 2019, there were 4,472 new VRE acquisition events at our hospital (569 clinical, 3,903 rectal screening). The overall VRE acquisition rate was 7.54 per 1,000 patient days (**Figure 1**). Incidence appeared to follow a seasonal pattern, with higher incidence in winter months and lower incidence in summer months. During the study period, we sequenced the genomes of 1,243 VRE isolates (891 rectal and 352 clinical) that were collected from 1096 unique patients. The sequenced isolates were predominantly *E. faecium* (n=1192, 96%) with the remaining belonging to *E. faecalis* (n=51, 4%). The *E. faecium* isolates were found to belong to 44 defined multi-locus sequence types (STs), with the most frequently sampled STs being ST17 (n=465, 37%), ST736 (n=174, 14%), and ST1471 (n=133, 11%) (**Table S1**).

**Figure 1.**
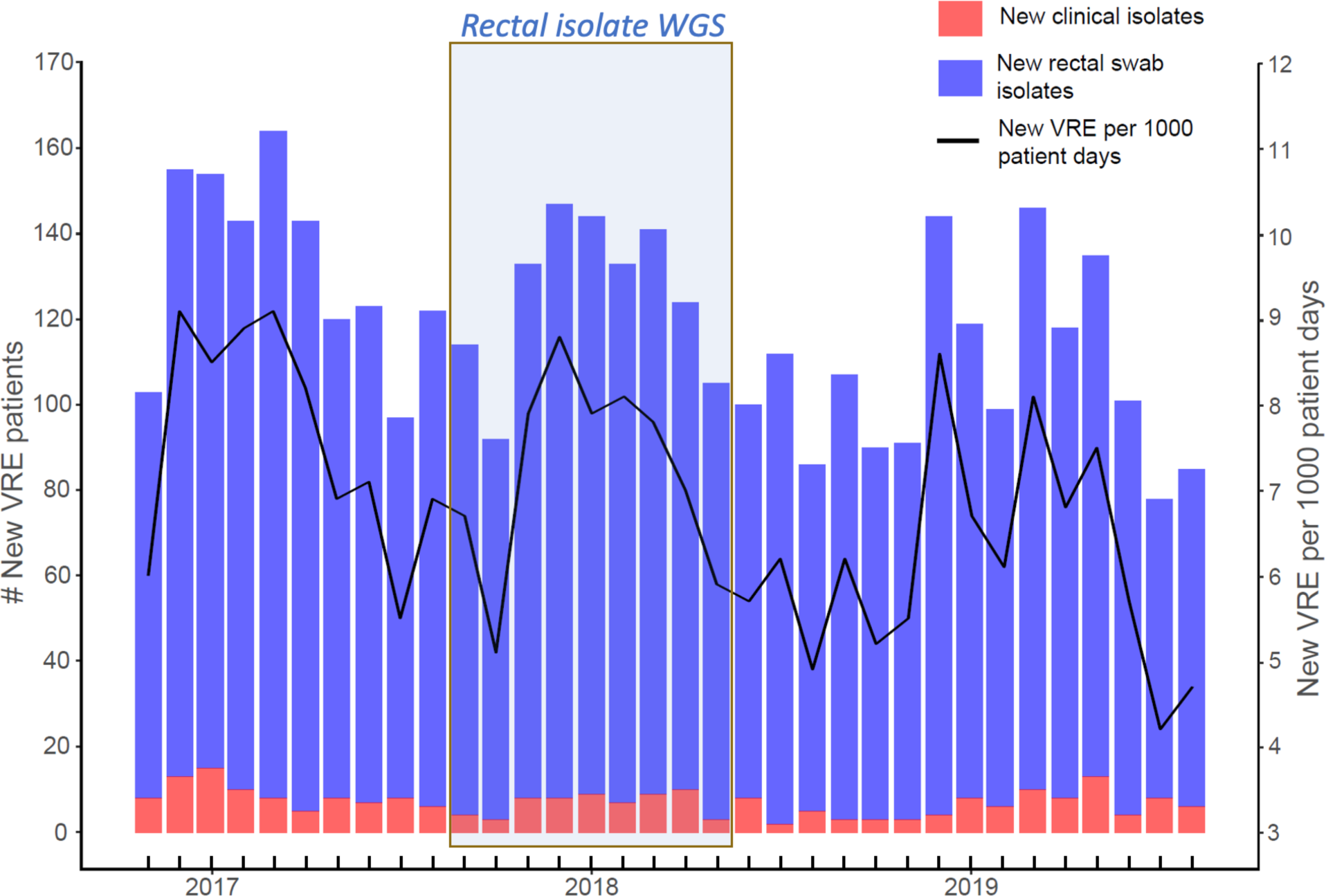
New VRE acquisition at a tertiary care hospital from November 2016 through August 2019. Newly identified VRE cases (clinical and rectal) per month for the study time period are shown. Year labels across the x-axis correspond to January of each year. Clinical isolates are shown in red, and rectal screening isolates are shown in blue. The black line indicates the rate of new VRE acquisition per 1000 patient days for each month.

### Genomic analysis of same-patient isolates

Of the 1096 patients sampled, 67 patients had multiple isolates, with 2-6 isolates collected from each patient. Within these patients, the number of days between culture dates of VRE isolates belonging to different STs was higher than that of isolates belonging to the same ST (mean: 148 days vs. 5 days, *P =* 0.0012), suggesting that some patients were colonized or infected with the same VRE strain, while others were colonized or infected with different strains (**Figure 2A**). When we compared isolates belonging to the same patient and same ST, the SNPs identified via split kmer analysis (SKA) showed higher variability compared to a reference-based SNP identification approach, supporting increased sensitivity in detecting SNP differences between closely related isolates (**Figure 2B**). As an extension to this analysis, we determined the rates of SNP accumulation over time for both SKA and reference-based SNPs using a SNP threshold of 20 SKA SNPs and 10 reference-based SNPs (upper whisker, 1.5 * IQR for each group, **Figure 2B**). SKA was more sensitive to determining SNP accumulation over time (*P* < 0.0001) as compared to SNPs identified using a reference-based approach, which did not show statistically significant differences in SNP accumulation over time (*P =* 0.2, **Figure 2C**).

**Figure 2.**
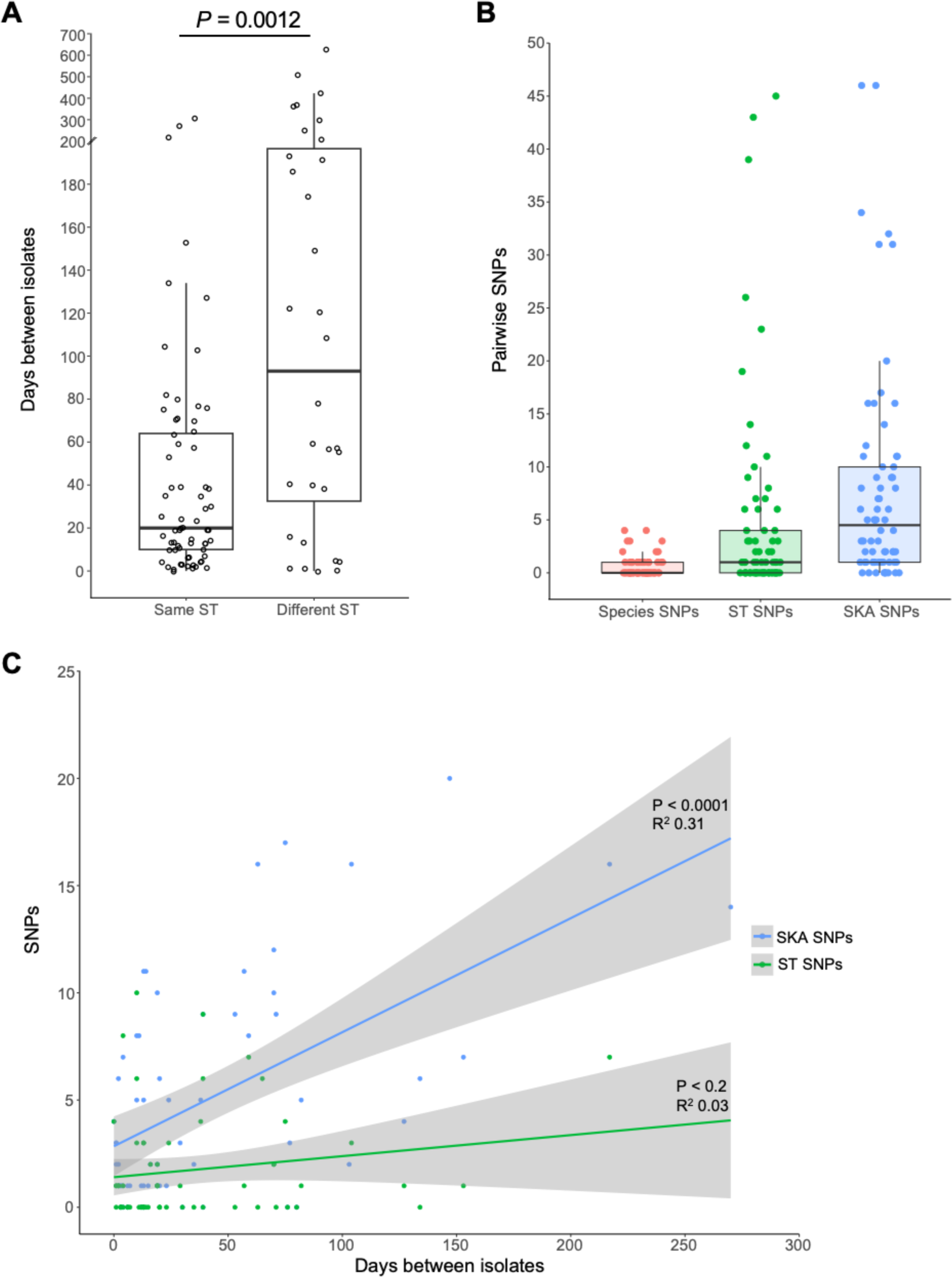
Genomic analysis of same-patient isolates. (A) Days between sampling of isolates belonging to the same sequence type (ST) versus different STs in patients that were repeatedly sampled. P-value is from a Mann Whitney U-test. (B) Pairwise single nucleotide polymorphisms (SNPs) between isolates collected from the same patient and belonging to the same ST, calculated using species SNPs (red), ST SNPs (green) and SKA SNPs (blue). (C) Pairwise SNPs versus days between samples for isolates collected from the same patient and belonging to the same ST, calculated with SKA SNPs (blue) or ST SNPS (green).

### Inclusion of rectal swab isolates increases linkage between clinical isolates

To identify groups of closely related isolates that might constitute putative transmission clusters, we performed hierarchical clustering using a cut-off of 20 SKA SNPs. Clustering of just clinical isolates identified 61 clusters, each containing 2-16 patients (median 2 patients per cluster). A total of 185 patient isolates (53%) clustered with at least one other isolate in this group. In contrast, clustering of all 1243 rectal and clinical isolates revealed 185 clusters, each containing 2-42 isolates (**Figure 3A, B**). A total of 233 clinical (66%) and 632 rectal swab isolates (71%) clustered with at least one other isolate in this group. The median SNP distance between clustered isolates was 8 SKA SNPs, which was slightly higher than the SNP distance between isolates collected from the same patient (median = 5 SKA SNPs) but was much lower than between isolates residing in different clusters (median = 120 SKA SNPs) (**Figure S2**). Compared with clustering of clinical isolates only, significantly more clinical isolates resided in clusters when rectal swab isolates were included (53% vs. 66%, *P* < 0.01). Additionally, the average cluster size was greater when rectal swab isolates were included (3.0 vs. 5.7 isolates per cluster, *P* < 0.05). Inclusion of rectal swab isolates also increased the linkage between clinical isolates compared to cluster analysis with clinical isolates only (*P* = 0.01) (**Figure 3C**).

**Figure 3.**
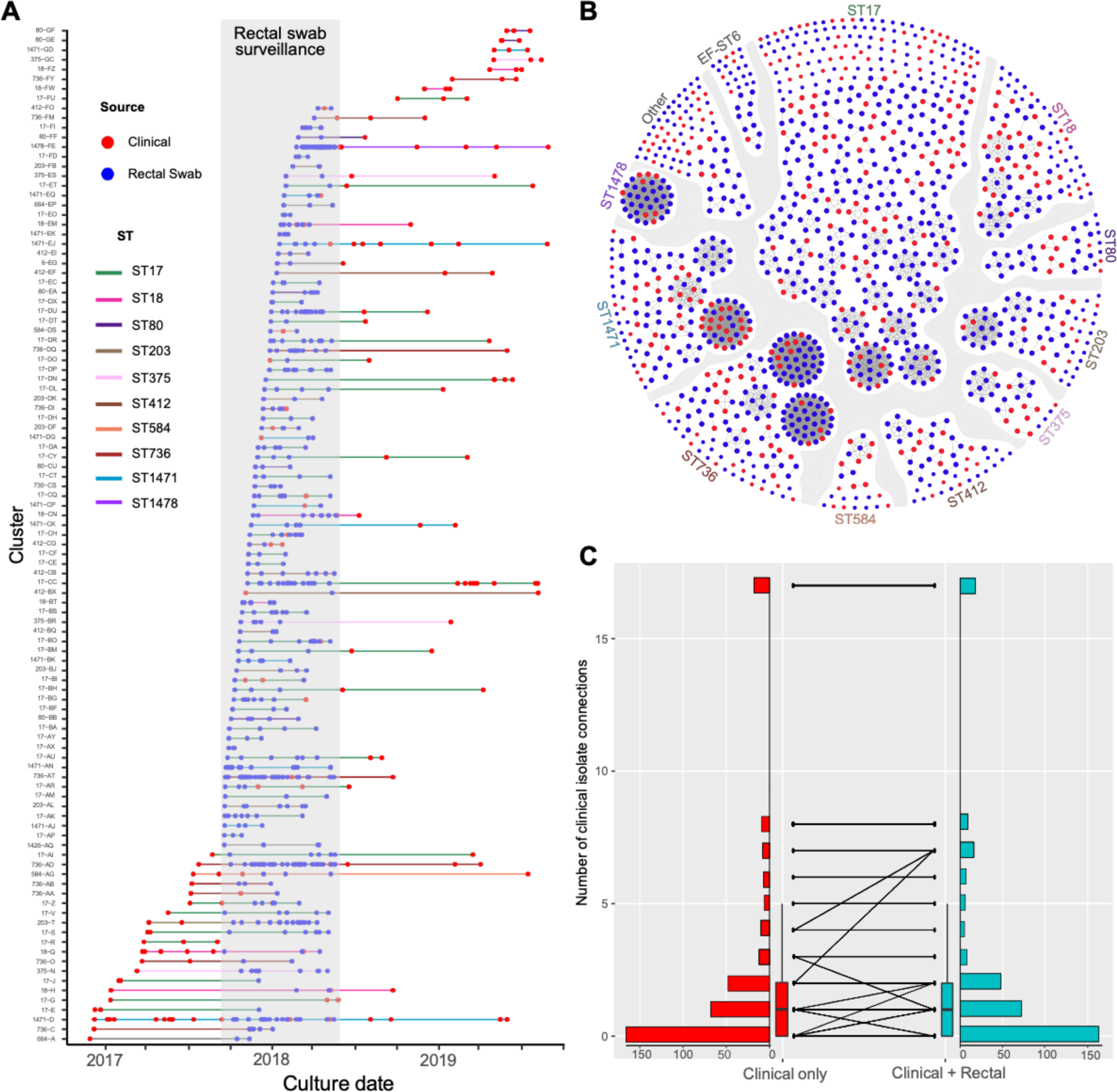
Inclusion of rectal swab isolates increases the number and size of genetically related VRE isolate clusters. (A) Waterfall timeline plot of isolate culture dates for 865 isolates belonging to 185 genetically related clusters. Isolates are colored by source and horizontal lines connecting isolates are colored by ST. The time period during which rectal swab isolates were collected is shaded in grey. (B) Cluster network diagram. All 1243 isolates are shown as nodes and are colored by collection source, with clinical isolates in red and rectal screening isolates in blue. Edges connect isolates within the same cluster. Isolates are grouped by MLST. All STs are *E. faecium*, except EF-ST6 which is *E. faecalis* ST6. (C) Raincloud plot showing cluster connections (i.e., edges in panel B) between clinical isolates when considering only clinical isolate clusters (red, left), versus clinical and rectal screening isolate clusters (teal, right).

### Inclusion of rectal swab isolates reveals increased transmission rates in healthcare settings

We performed a geo-temporal analysis to identify potential sources of transmission in our hospital (**Figure S1**). Considering clusters containing both clinical and rectal swab isolates, geo-temporal analysis revealed potential sources of transmission for 271 (31.3%) isolates (224 rectal, 47 clinical) (**Table S2**). Considering clinical isolates alone, the same analysis revealed possible sources of transmission for only 33 (17.8%) clustered clinical isolates. A general intensive care unit was the unit with the highest number of infections with potential geo-temporal links in the combined clinical and rectal dataset (55 infections) as well as in the clinical-only dataset (5 infections) (**Figure 4**). During the rectal screening time frame, we observed a higher transmission rate (defined as the number of days between related isolates) than during the clinical-only time frame (*P* < 0.0001) (**Figure S3**). Among the 120 clinically clustered isolates that were closely related to prior rectal isolates, VRE was associated with 11 (9.2%) 30-day readmissions, 21 (17.5%) ICU admissions, and 16 (13.3%) deaths (**Table S3**). Only 42 (35.0%) of these infections were reported as healthcare-associated.

**Figure 4.**
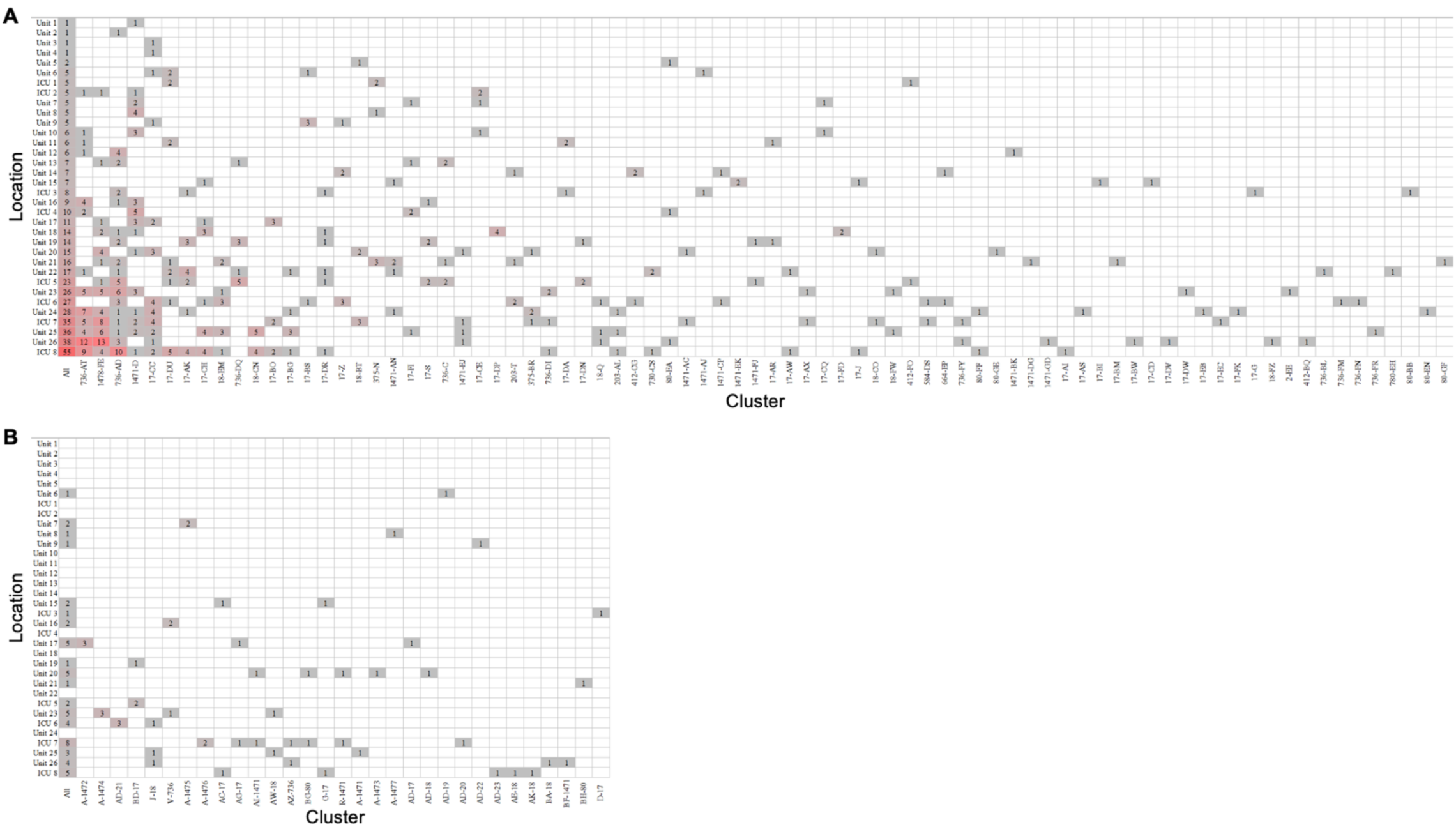
Hospital units associated with VRE transmission. (A) Heatmap showing geo-temporal transmission links between isolates in each cluster, considering both clinical and rectal screening isolates. (B) Heatmap showing geo-temporal transmission links between clustered clinical isolates only. Counts within each cell represents the total number of isolates in the cluster (vertical columns) with possible unit transmission (horizontal rows). Leftmost columns show the total counts across all columns.

## DISCUSSION

In this study, we examined the genomic epidemiology of VRE at our hospital spanning a period of 34 months. We observed high rates of VRE transmission, which was particularly evident when focusing on rectal screening isolates during a nine-month nested period. Our data highlight the complexities of understanding transmission of VRE in healthcare settings, but also demonstrate the usefulness of WGS surveillance to elucidate and better understand these patterns.

Prior studies have shown that VRE is endemic within healthcare systems worldwide and spreads through the dissemination of polyclonal lineages. Prior studies have reported clonal domination of vancomycin-resistant *E. faecium* lineages ST17, ST80, ST117, and ST1478, all of which belong to the hospital-adapted clonal complex CC-17.^29–33^ Such lineages have been responsible for widespread transmission events within healthcare systems. Similarly, we have identified the presence of these global, problematic lineages at high incidence within our own hospital. Prior studies have characterized healthcare-associated VRE transmission rates ranging from 60-80%.^9,34^ Consistent with these prior reports, we found that 70% of VRE isolates in our study were genetically linked to one another when we considered both clinical and rectal swab isolates, however this proportion fell to only 53% when we considered clinical isolates only. Together these findings suggest that the inclusion of rectal swabs increases clustering of related isolates, and may improve both within and between patient lineage tracking.

Data obtained from rectal swabs, even over a relatively short duration of nine months, enhanced our understanding of the transmission dynamics of clinical VRE infections. Performing WGS surveillance on clinical isolates alone missed large numbers of rectal transmission events and also missed likely sources of transmission of clinical infections. Without inclusion of rectal swabs, interventions directed as clinical transmission alone may be misdirected. This highlights the potential of systematic surveillance in providing key insights into the behavior and spread of VRE, which is continuing to emerge as more common and pathogenic.^35^ Within our study, contact precautions were used, however the efficacy of contact precautions in reducing VRE transmission has been highly debated, with discontinuation showing no associated increase in HAIs.^36^ With 70% of our VRE isolates genetically related to one another, our data question the effectiveness of contact precautions in preventing VRE transmission.

We found that VRE infections associated with readmissions (9.2%) and deaths (13.3%) could be genetically linked with prior rectal screening isolates. These proportions indicate the role of rectal colonization as a transmission source that contributes to subsequent, severe clinical outcomes. Targeting of rectal colonization with infection prevention measures informed by sequencing-based surveillance could address this issue by potentially curbing onward VRE transmission and decreasing the rates of poor outcomes due to VRE infection.

The data presented in this study, along with findings from other research highlighting the advantages of WGS surveillance in hospital settings, strongly advocate for the routine, prospective use of WGS surveillance.^22,37^ This approach could enhance patient safety and contribute to a more comprehensive understanding of pathogen evolution and transmission. As hospitals navigate the complexities of HAIs, such as those caused by VRE colonization and infection, the integration of innovative genomic technologies like WGS into routine diagnostic protocols could make a substantial impact. The ability of WGS to quantify genetic differences within and between bacterial strains can help accurately identify sources and patterns of pathogen spread, providing key insights necessary to design effective control strategies.

Our study has several limitations. First, we only sequenced VRE isolates from infections that were potentially healthcare-associated. However, this would only underestimate additional related isolates that were introduced from community infections. Second, we only sequenced rectal isolates during a nine-month nested period. Regardless, we still found a high percentage of genetic relatedness among these isolates. Third, we only considered geo-temporal analysis among clustered isolates and did not consider common procedures or common healthcare personnel. Fourth, clusters of related isolates may represent common circulating community strains. However, we did find geo-temporal links connecting many of these infections within our hospital. Fifth, we only had access to the VRE isolates collected during the study period. Some patients may have had prior VRE colonization or infection, which may have miscategorized their study infections as new acquisitions.

In summary, we describe the genomic epidemiology of VRE in a hospital setting over a nearly three-year period. The high rates of VRE transmission observed, especially among rectal screening isolates, underscore the crucial role of systematic surveillance in understanding and mitigating the spread of VRE. The real-time collection and WGS surveillance of VRE isolates into hospital practice could potentially aid infection prevention efforts to prevent future transmission and clinical infections, thus enhancing patient safety.

## Data Availability

All data produced in the present study are available upon reasonable request to the authors

## ACKNOWLEDGMENTS

We gratefully acknowledge Jane Marsh, Vaughn Cooper, and Elise Martin for their expert advice and assistance throughout the course of this study.

## Author Contributions

AJS: Conceptualization, Methodology, Analysis, Data Curation, Writing - Original Draft, Writing - Review & Editing, Visualization

VRS: Analysis, Data Curation, Writing - Review & Editing, Visualization

EGM: Methodology, Analysis, Data Curation, Writing - Review & Editing, Visualization

MPG: Data Curation

EE: Writing - Review & Editing

JC: Data Curation

KDW: Data Curation

GMS: Conceptualization, Writing - Review & Editing

LLP: Writing - Review & Editing

LHH: Resources, Writing - Review & Editing, Funding acquisition

DVT: Conceptualization, Methodology, Analysis, Resources, Data Curation, Writing - Review & Editing, Visualization, Supervision, Project administration, Funding acquisition

## STUDY FUNDING

This work was funded in part by the National Institutes of Health (NIH) through grants R01AI127472 to LHH and R01AI165519 to DVT. The NIH played no role in data collection, analysis, or interpretation; study design; writing of the manuscript; or decision to submit for publication.

## Declaration of Interests

None

## SUPPLEMENTAL INFORMATION

**Table S1. List of isolates in the dataset, including isolate ID, year of collection, species, source, cluster, and ST.**

**Table S2.** Geo-temporal links between clinical versus clinical and rectal isolates.

|  | Isolates Related |  |  | Isolates with Geo-temporal Links (%) |  |  |
| --- | --- | --- | --- | --- | --- | --- |
|  | Clinical Isolates | Rectal Isolates | Total Isolates | Clinical Isolates | Rectal Isolates | Total |
| Clinical-only | 185 | NA | 185 | 33 (17.8) | NA | 33 (17.8) |
| Clinical + Rectal | 233 | 632 | 865 | 47 (20.2) | 224 (35.4) | 271 (31.3) |

**Table S3.**
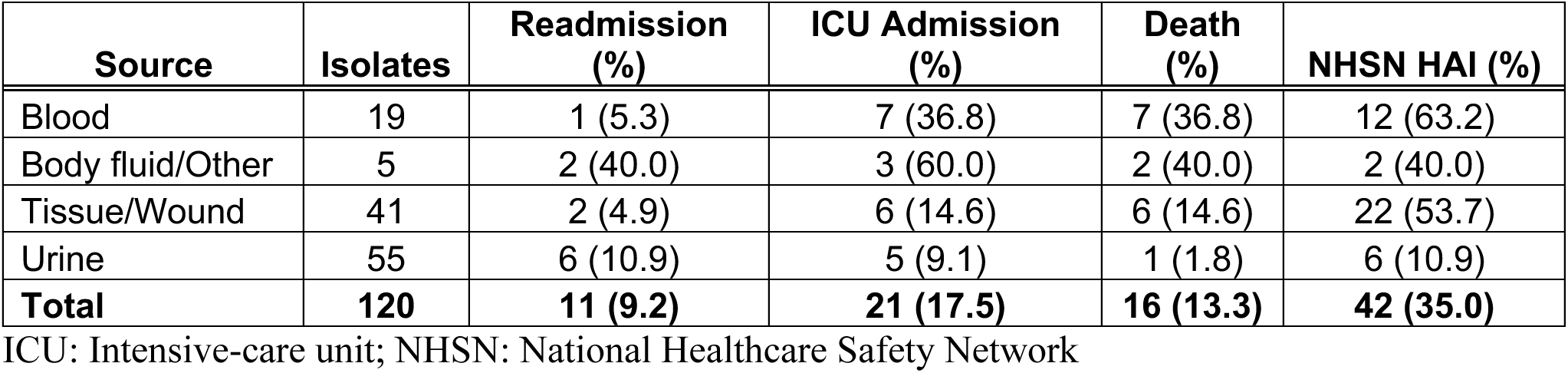
30-day outcomes of 120 VRE clinical isolates that clustered with a prior rectal isolate.

| Source | Isolates | Readmission (%) | ICU Admission (%) | Death (%) | NHSN HAI (%) |
| --- | --- | --- | --- | --- | --- |
| Blood | 19 | 1 (5.3) | 7 (36.8) | 7 (36.8) | 12 (63.2) |
| Body fluid/Other | 5 | 2 (40.0) | 3 (60.0) | 2 (40.0) | 2 (40.0) |
| Tissue/Wound | 41 | 2 (4.9) | 6 (14.6) | 6 (14.6) | 22 (53.7) |
| Urine | 55 | 6 (10.9) | 5 (9.1) | 1 (1.8) | 6 (10.9) |
| **Total** | **120** | **11 (9.2)** | **21 (17.5)** | **16 (13.3)** | **42 (35.0)** |

ICU: Intensive-care unit; NHSN: National Healthcare Safety Network

**Figure S1.**
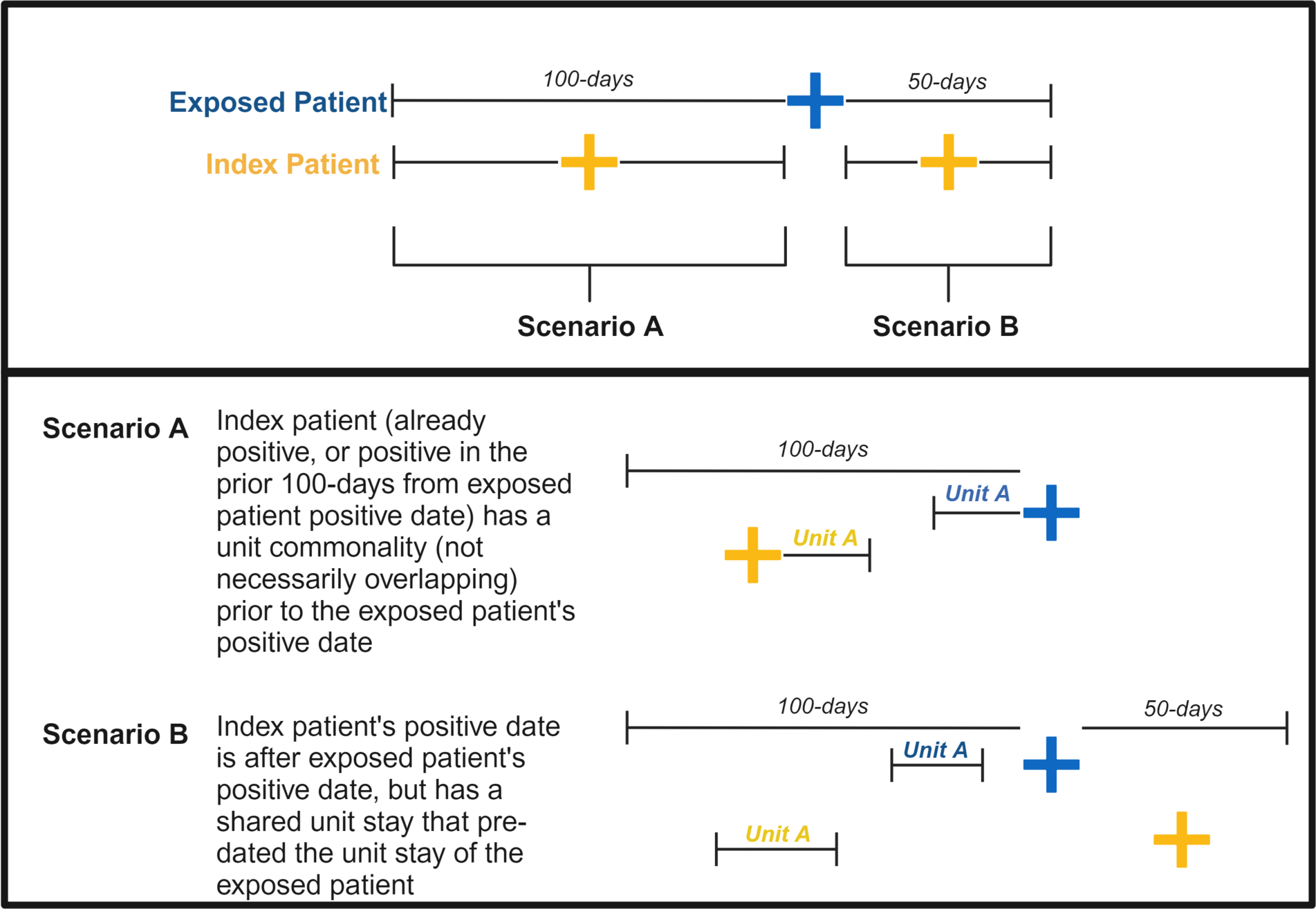
Geo-temporal transmission analysis approach.

**Figure S2.**
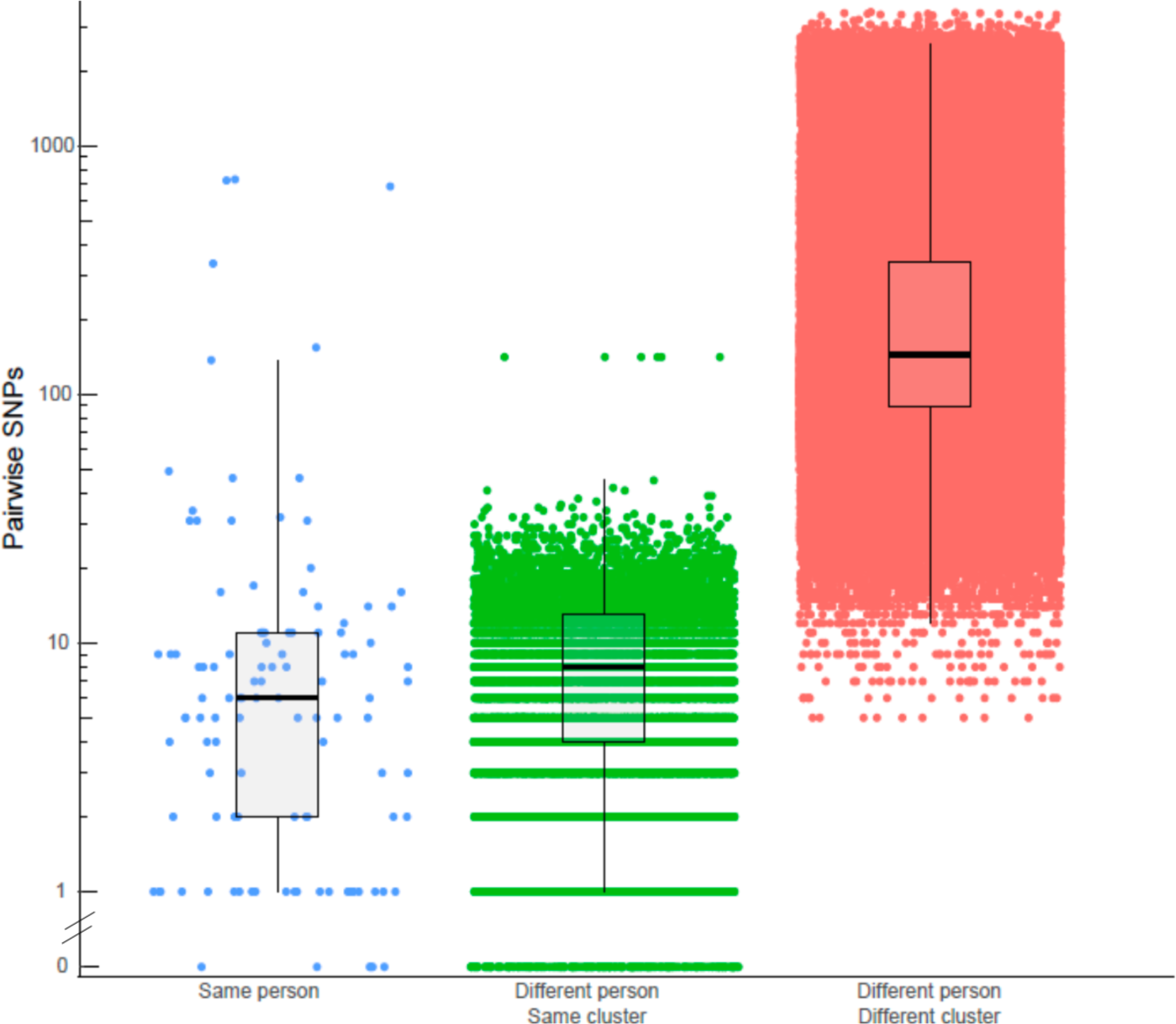
SNP distance comparisons. Pairwise SKA SNPs were calculated for all same-person isolates (blue), same-cluster isolates (green), and different person/different cluster isolates (red).

**Figure S3.**
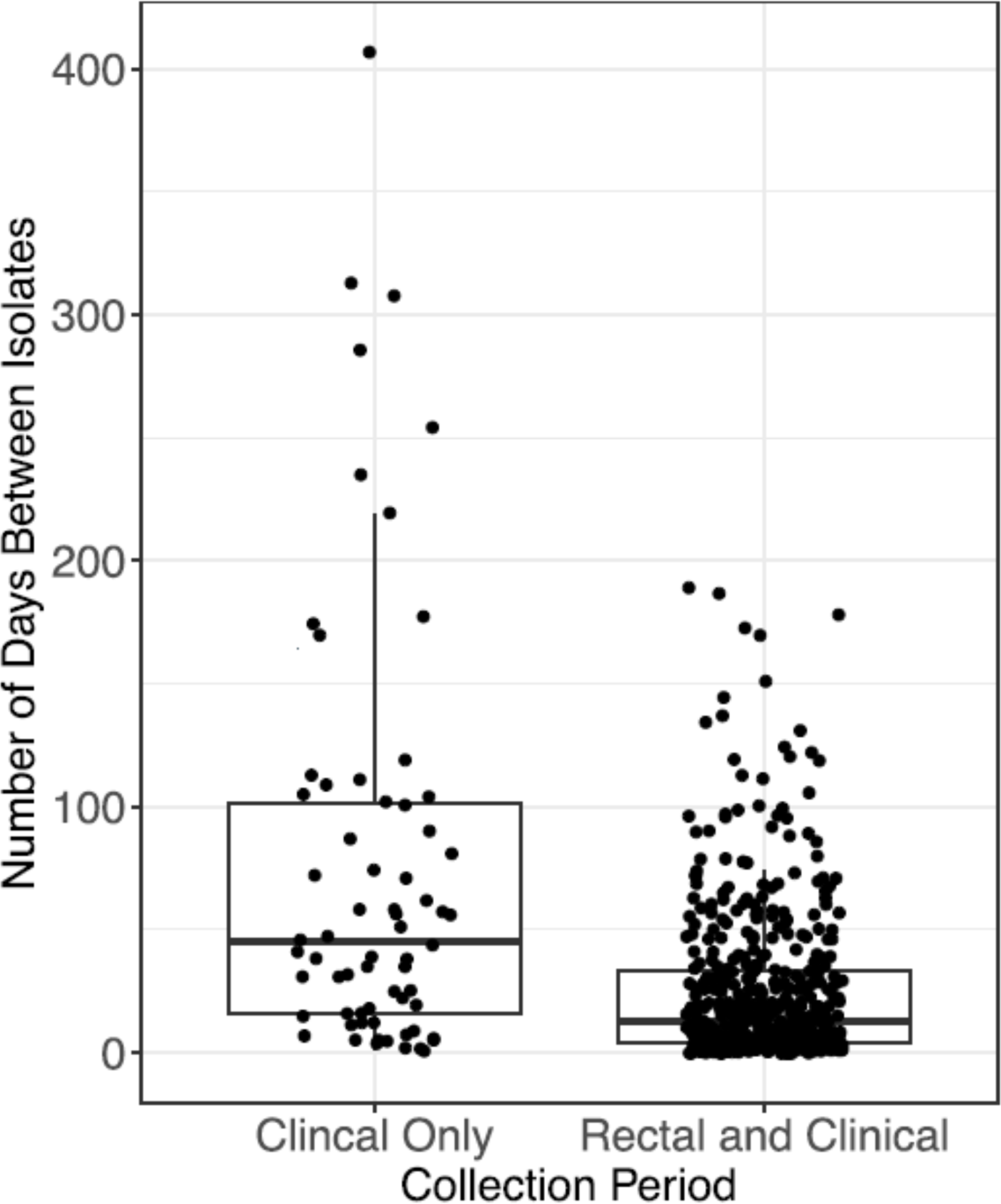
Transmission rates in the non-rectal screening versus clinical and rectal screening time periods. Transmission rate was defined as the number of days between collection of related isolates.

## Notes

### Competing Interest Statement

The authors have declared no competing interest.

### Author Declarations

Ethics approval was obtained from the University of Pittsburgh Institutional Review Board (Protocol STUDY21040126).

## REFERENCES

1. VRE in Healthcare Settings | HAI | CDC. Published November 6, 2019. Accessed February 8, 2024. https://www.cdc.gov/hai/organisms/vre/vre.html

2. Eichel V, Klein S, Bootsveld C, et al. Challenges in interpretation of WGS and epidemiological data to investigate nosocomial transmission of vancomycin-resistant Enterococcus faecium in an endemic region: incorporation of patient movement network and admission screening. J Antimicrob Chemother. 2020;75(7):1716–1721. doi:10.1093/jac/dkaa122

3. Lankford MG, Collins S, Youngberg L, Rooney DM, Warren JR, Noskin GA. Assessment of materials commonly utilized in health care: implications for bacterial survival and transmission. Am J Infect Control. 2006;34(5):258–263. doi:10.1016/j.ajic.2005.10.008

4. O’Driscoll T, Crank CW. Vancomycin-resistant enterococcal infections: epidemiology, clinical manifestations, and optimal management. Infect Drug Resist. 2015;8:217–230. doi:10.2147/IDR.S54125

5. Kramer TS, Remschmidt C, Werner S, et al. The importance of adjusting for enterococcus species when assessing the burden of vancomycin resistance: a cohort study including over 1000 cases of enterococcal bloodstream infections. Antimicrob Resist Infect Control. 2018;7:133. doi:10.1186/s13756-018-0419-9

6. Contreras GA, Munita JM, Simar S, et al. Contemporary Clinical and Molecular Epidemiology of Vancomycin-Resistant Enterococcal Bacteremia: A Prospective Multicenter Cohort Study (VENOUS I). Open Forum Infect Dis. 2022;9(3):ofab616. doi:10.1093/ofid/ofab616

7. Liese J, Schüle L, Oberhettinger P, et al. Expansion of Vancomycin-Resistant Enterococcus faecium in an Academic Tertiary Hospital in Southwest Germany: a Large-Scale Whole-Genome-Based Outbreak Investigation. Antimicrob Agents Chemother. 2019;63(5):e01978–18. doi:10.1128/AAC.01978-18

8. Arias CA, Murray BE. The rise of the Enterococcus: beyond vancomycin resistance. Nat Rev Microbiol. 2012;10(4):266–278. doi:10.1038/nrmicro2761

9. Gouliouris T, Coll F, Ludden C, et al. Quantifying acquisition and transmission of Enterococcus faecium using genomic surveillance. Nat Microbiol. 2021;6(1):103–111. doi:10.1038/s41564-020-00806-7

10. Marom R, Mandel D, Haham A, et al. A silent outbreak of vancomycin-resistant Enterococcus faecium in a neonatal intensive care unit. Antimicrob Resist Infect Control. 2020;9(1):87. doi:10.1186/s13756-020-00755-0

11. Leong KWC, Cooley LA, Anderson TL, et al. Emergence of Vancomycin-Resistant Enterococcus faecium at an Australian Hospital: A Whole Genome Sequencing Analysis. Sci Rep. 2018;8(1):6274. doi:10.1038/s41598-018-24614-6

12. García Martínez de Artola D, Castro B, Ramos MJ, Díaz Cuevas Z, Lakhwani S, Lecuona M. Outbreak of vancomycin-resistant enterococcus on a haematology ward: management and control. J Infect Prev. 2017;18(3):149–153. doi:10.1177/1757177416687832

13. Andersson P, Beckingham W, Gorrie CL, et al. Vancomycin-resistant Enterococcus (VRE) outbreak in a neonatal intensive care unit and special care nursery at a tertiary-care hospital in Australia-A retrospective case-control study. Infect Control Hosp Epidemiol. 2019;40(5):551–558. doi:10.1017/ice.2019.41

14. Frakking FNJ, Bril WS, Sinnige JC, et al. Recommendations for the successful control of a large outbreak of vancomycin-resistant Enterococcus faecium in a non-endemic hospital setting. J Hosp Infect. 2018;100(4):e216–e225. doi:10.1016/j.jhin.2018.02.016

15. Piezzi V, Wassilew N, Atkinson A, et al. Nosocomial outbreak of vancomycin-resistant Enterococcus faecium (VRE) ST796, Switzerland, 2017 to 2020. Euro Surveill. 2022;27(48):2200285. doi:10.2807/1560-7917.ES.2022.27.48.2200285

16. Abdelbary MHH, Senn L, Greub G, Chaillou G, Moulin E, Blanc DS. Whole-genome sequencing revealed independent emergence of vancomycin-resistant Enterococcus faecium causing sequential outbreaks over 3 years in a tertiary care hospital. Eur J Clin Microbiol Infect Dis. 2019;38(6):1163–1170. doi:10.1007/s10096-019-03524-z

17. Tang P, Croxen MA, Hasan MR, Hsiao WWL, Hoang LM. Infection control in the new age of genomic epidemiology. Am J Infect Control. 2017;45(2):170–179. doi:10.1016/j.ajic.2016.05.015

18. Udaondo Z, Abram K, Kothari A, Jun SR. Top-Down Genomic Surveillance Approach To Investigate the Genomic Epidemiology and Antibiotic Resistance Patterns of Enterococcus faecium Detected in Cancer Patients in Arkansas. Microbiol Spectr. 2023;11(3):e0490122. doi:10.1128/spectrum.04901-22

19. Permana B, Harris PNA, Runnegar N, et al. Using Genomics To Investigate an Outbreak of Vancomycin-Resistant Enterococcus faecium ST78 at a Large Tertiary Hospital in Queensland. Microbiol Spectr. 2023;11(3):e0420422. doi:10.1128/spectrum.04204-22

20. Raven KE, Gouliouris T, Brodrick H, et al. Complex Routes of Nosocomial Vancomycin-Resistant Enterococcus faecium Transmission Revealed by Genome Sequencing. Clin Infect Dis. 2017;64(7):886–893. doi:10.1093/cid/ciw872

21. Brodrick HJ, Raven KE, Harrison EM, et al. Whole-genome sequencing reveals transmission of vancomycin-resistant Enterococcus faecium in a healthcare network. Genome Med. 2016;8(1):4. doi:10.1186/s13073-015-0259-7

22. Sundermann AJ, Chen J, Kumar P, et al. Whole-Genome Sequencing Surveillance and Machine Learning of the Electronic Health Record for Enhanced Healthcare Outbreak Detection. Clinical Infectious Diseases. 2022;75(3):476–482. doi:10.1093/cid/ciab946

23. Prjibelski A, Antipov D, Meleshko D, Lapidus A, Korobeynikov A. Using SPAdes De Novo Assembler. Curr Protoc Bioinformatics. 2020;70(1):e102. doi:10.1002/cpbi.102

24. Jolley KA, Bray JE, Maiden MCJ. Open-access bacterial population genomics: BIGSdb software, the PubMLST.org website and their applications. Wellcome Open Res. 2018;3:124. doi:10.12688/wellcomeopenres.14826.1

25. Harris SR. SKA: Split Kmer Analysis Toolkit for Bacterial Genomic Epidemiology. Published online October 25, 2018:453142. doi:10.1101/453142

26. Sundermann AJ, Griffith MP, Srinivasa VR, et al. Prolonged bacterial carriage and hospital transmission detected by whole genome sequencing surveillance. Antimicrobial Stewardship & Healthcare Epidemiology. 2024;4(1):e11. doi:10.1017/ash.2024.4

27. NHSN | CDC. Published January 4, 2024. Accessed February 8, 2024. https://www.cdc.gov/nhsn/index.html

28. Sundermann AJ, Penzelik J, Ayres A, Snyder GM, Harrison LH. Sensitivity of National Healthcare Safety Network definitions to capture healthcare-associated transmission identified by whole-genome sequencing surveillance. Infect Control Hosp Epidemiol. 2023;44(10):1663–1665. doi:10.1017/ice.2023.52

29. McCracken M, Mitchell R, Smith S, et al. Emergence of pstS-Null Vancomycin-Resistant Enterococcus faecium Clone ST1478, Canada, 2013-2018. Emerg Infect Dis. 2020;26(9):2247–2250. doi:10.3201/eid2609.201576

30. Egan SA, Kavanagh NL, Shore AC, et al. Genomic analysis of 600 vancomycin-resistant Enterococcus faecium reveals a high prevalence of ST80 and spread of similar vanA regions via IS1216E and plasmid transfer in diverse genetic lineages in Ireland. J Antimicrob Chemother. 2022;77(2):320–330. doi:10.1093/jac/dkab393

31. Fang H, Fröding I, Ullberg M, Giske CG. Genomic analysis revealed distinct transmission clusters of vancomycin-resistant Enterococcus faecium ST80 in Stockholm, Sweden. J Hosp Infect. 2021;107:12–15. doi:10.1016/j.jhin.2020.10.019

32. Pinholt M, Larner-Svensson H, Littauer P, et al. Multiple hospital outbreaks of vanA Enterococcus faecium in Denmark, 2012-13, investigated by WGS, MLST and PFGE. J Antimicrob Chemother. 2015;70(9):2474–2482. doi:10.1093/jac/dkv142

33. Jozefíková A, Valček A, Šoltys K, Nováková E, Bujdáková H. Persistence and multi-ward dissemination of vancomycin-resistant Enterococcus faecium ST17 clone in hospital settings in Slovakia 2017-2020. Int J Antimicrob Agents. 2022;59(4):106561. doi:10.1016/j.ijantimicag.2022.106561

34. Sherry NL, Gorrie CL, Kwong JC, et al. Multi-site implementation of whole genome sequencing for hospital infection control: A prospective genomic epidemiological analysis. Lancet Reg Health West Pac. 2022;23:100446. doi:10.1016/j.lanwpc.2022.100446

35. Rödenbeck M, Ayobami O, Eckmanns T, Pletz MW, Bleidorn J, Markwart R. Clinical epidemiology and case fatality due to antimicrobial resistance in Germany: a systematic review and meta-analysis, 1 January 2010 to 31 December 2021. Euro Surveill. 2023;28(20):2200672. doi:10.2807/1560-7917.ES.2023.28.20.2200672

36. Martin EM, Colaianne B, Bridge C, et al. Discontinuing MRSA and VRE contact precautions: Defining hospital characteristics and infection prevention practices predicting safe de-escalation. Infect Control Hosp Epidemiol. 2022;43(11):1595–1602. doi:10.1017/ice.2021.457

37. Sundermann AJ, Chen J, Miller JK, et al. Whole-genome sequencing surveillance and machine learning for healthcare outbreak detection and investigation: A systematic review and summary. Antimicrob Steward Healthc Epidemiol. 2022;2(1):e91. doi:10.1017/ash.2021.241

